# Etiologic Subtypes of Ischemic Stroke in SARS-COV-2 Virus patients in a cohort of New York City hospitals

**DOI:** 10.1101/2020.05.03.20077206

**Authors:** Ketevan Berekashvili, Adam A. Dmytriw, Volodomyr Vulkanov, Shashank Agarwal, Amit Khaneja, Thambirajah Nandakumar, Jeremy Liff, David Turkel-Parrella, Jeffrey Farkas, Ting Zhou, Jennifer A. Frontera, David E. Kahn, Sun Kim, Kelley A. Humbert, Matthew D. Sanger, Shadi Yaghi, Aaron S. Lord, Karthikeyan M. Arcot, Ambooj Tiwari

## Abstract

**Objective:** To describe the ischemic stroke subtypes related to COVID-19 in a cohort of NYC hospitals and explore their etiopathogenesis.

**Background:** Extra-pulmonary involvement of COVID-19 has been reported in the hepatic, renal and hematological systems. Most neurological manifestations are non-focal, but few have reported the characteristics of ischemic strokes or investigated its pathophysiology.

**Methods:** Data were collected prospectively from March 15 to April 15, 2020 from four centers in New York City to review possible ischemic stroke types seen in COVID-19 positive patients. Patient presentation, demographics, other related vascular risk factors, associated laboratory and coagulation markers, as well as imaging and outcomes for consecutive stroke patients positive for SARS-COV2 infection over the period studied were collected.

**Results:** In our study, the age range of patients was 25-75 with no significant male preponderance. The median age of LVO patients was 48. Stroke was the presenting and hospitalizing event in 70%. One fifth of patients did not have common risk factors for ischemic stroke and none had atrial fibrillation, coronary or cerebrovascular disease, or were smokers. Half had a poor outcome with 40% ending in mortality (60% in LVO group) and one in a critical condition due ARDS. All had high neutrophil/lymphocyte ratio except one who demonstrated some neurological recovery. D-dimer levels showed mild to severe elevation when collected. None of the LVO cases had known cardiac risk factors but two out of five were found to have cardiac abnormalities during their hospitalization. All LVOs had hypercoagulable lab markers especially elevated D-dimer and/or fibrinogen. The LVO patients were younger and sicker with a median age of 46 and mean NIHSS of 24 as opposed to non-LVOs with a median age of 62 and mean NIHSS of 6 respectively.

**Conclusion:** COVID-19 related ischemic events can be small vessel, branch emboli or large vessel occlusions. The latter is often associated with either a hypercoagulable state or cardio-embolism. Patient outcomes were worse when multi-organ or pulmonary system failure prevailed.

## Introduction

The novel Coronavirus outbreak came to the fore in December 2019. Several features have been common manifestations of this disease: a primarily lower respiratory tract illness, a severe form that is more common in people with underlying diseases and higher mortality/case fatality in older populations. (1–4) A particularly virulent form of immunological response, the cytokine storm syndrome especially seems to affect these vulnerable subgroups. (4–6) This syndrome is also often associated with extra-pulmonary complications of COVID-19. (5,7)

In the last three months, multiple case reports have highlighted the extra-pulmonary complications of the disease. (7–10) A prominent subgroup of these include thromboembolic complications. (9–11). They have also been often associated with lab markers suggesting an underlying inflammatory and hypercoagulable condition. (9,11,12) To this end, the specter of a COVID-19-specific coagulopathy has been raised, which is both inflammatory and consumptive, different from traditional disseminated intravascular coagulopathy (9).

Most neurological complications have focused on non-focal presentations like headaches, encephalopathy and skeletal muscle injury. (8,13) While some authors have mentioned the possibility of a form of meningoencephalitis, viral causation of neurological diseases have been mentioned less often. (14,15) Reports of patients presenting with focal signs secondary to ischemic strokes have been scant. (8,16)

The objective of this study was to prospectively study COVID-19 presenting as focal neurological disease caused by arterial thromboembolism to the intracranial circulation. We also wanted to better describe the different ischemic stroke subtypes seen in patients with SARS-COV2 infection especially with the view to assess its unique features seen in the context of COVID-19.

## Methods

Retrospective case review protocol approval was obtained from the Institutional Review Boards at NYU School of Medicine, Brookdale Hospital University Medical Center, Jamaica Medical Center and Richmond University Medical Centers respectively. A retrospective analysis of the prospectively maintained stroke databases at these institutions between the period of March 15 to April 15, 2020 was performed. The period coincided with widespread testing in the hospitals where these patients were admitted. Patients who tested positive with SARS-COV-2 PCR were selected (Cobas SARS-COV-2 real time RT-PCR under EUA performed & Abbott Real-time SARS-CoV-2 PCR assay using M2000 platform). Only patients with ischemic strokes were selected. We excluded other neurovascular events like intracerebral hemorrhage, subarachnoid hemorrhage or venous thrombosis for the purpose of this analysis. The results from four NYC hospitals were pooled including two in Brooklyn, one in Queens and one in Staten Island.

Patients with both TIA and ischemic strokes were selected for this purpose. The clinical definition of an acute onset of a focal deficit to characterize an ischemic event was used. All patients underwent rapid imaging with CT and CT angiogram to confirm presence of a parenchymal lesions and/or vascular occlusion. Patients with ASPECTS (Alberta Stroke Program Early CT Score) of 6 ore greater were selected for mechanical thrombectomy. Parenchymal imaging with CT or MRI was also performed in the post-acute period to define the areas of infarction.

Demographics including age and sex as well as history of other vascular risk factors like diabetes, hypertension, smoking and atrial fibrillation were collected. COVID-19 related history including other possible underlying conditions (e.g. Asthma, COPD, immunocompromised status), timeline from first symptoms (when available), pulmonary and extra-pulmonary symptoms as well as relevant laboratory information was also collected. Stroke variables included type of clinical presentation, parenchymal and vascular imaging findings, Trial of Org 10172 in Acute Stroke Treatment (TOAST) criteria for etiology, cardiac history for LVO cases, time and type of treatment from index event and pre-procedural neurological (NIHSS). Outcome variables included the following: discharge mRS, mortality as well as final imaging results. Descriptive statistics were utilized as appropriate.

## Results

Ten patients were included in this case series from four New York City hospitals: Richmond University Medical Center-Staten Island (1 patient), NYU Langone Health-Brooklyn (2 patients), Jamaica Hospital Medical Center (2 patients) and Brookdale Hospital Medical Center (5 patients).

Age of patients ranged between 27 and 75 years. About 80% of the patients had pre-existing conditions, the most common being diabetes (70%) and hypertension (60%). While some patients had comorbidities, no patient had preexisting atrial fibrillation, coronary disease, or cerebrovascular disease. One of the patients developed atrial fibrillation during their course in hospital. The majority of patients encountered (80%) presented to the emergency room (ER) with chief complaints of neurological deficits rather than respiratory symptoms (Table 1). Non-large vessel occlusions comprised lacunar infarcts, branch emboli or transient ischemic event (Table 2). Of note, only two patients (20%) presented within the 4.5 hour window for intravenous thrombolysis. Six (50%) presented more than 12 hours from last known normal and 1 presented more than 24 hours from last known normal. Nine patients (90%) were racial minorities and more than half (60%) were African-American.

**Table 1:** Characteristics of Patients with Large Vessel Occlusion

| Characteristic | Patient 1 | Patient 2 | Patient 3 | Patient 4 | Patient 5 |
| --- | --- | --- | --- | --- | --- |
| Demographics |  |  |  |  |  |
| Age (years) | 50-60 | 70-80 | 30-40 | 40-50 | 20-30 |
| Race | Asian | Hispanic | African American | African American | Asian |
| Sex | Male | Female | Male | Male | Male |
| Initial Findings |  |  |  |  |  |
| Medical history | DM, HTN | DM, HTN, CVA | DM, HTN | None | None |
| Symptoms at disease onset | Fever, cough, hypoxia, diarrhea | Neurological, Fever, hypoxia | Neurological, cough | Neurological Hypoxia, coma | Fever, Cough |
| Stroke syndrome | Right MCA | Right MCA | Left MCA | Posterior circulation | Left MCA |
| NIHSS | 24 | 13 | 26 | 31 | 18 |
| Cardiac history | NSR, | AFib + RVR, | NSR, | NSR, | NSR, |
|  | Normal EF | Normal EF | LVEF 26% | Normal EF | Normal EF |
| Imaging |  |  |  |  |  |
| CXR | Patchy parenchymal opacities bilateral | Patchy parenchymal opacities bilateral | Left small hazy opacity | Hazy opacities bilateral lower fields | bilateral patchy infiltrate |
| CT head | ASPECTS 5 | ASPECT 8 | ASPECTS 6 | Early pontine infarcts | ASPECTS 9 |
| CTA Head/Neck | Right Supra-clinoid ICA T occlusion | Right M1 occlusion | Left distal M1 occlusion | Proximal basilar occlusion | Left M1 |
| Laboratory |  |  |  |  |  |
| D-Dimer | 10,000 | 3247 | 769 | >10,000 | 4964 |
| NLR | 4.2 | 5.2 | 1.7 | 17.4 | 8 |
| Fibrinogen | 235 | 634 | 542 | 316 | 970 |
| Ferritin | 588 | 24 | 207 | 3746 | 1350 |
| Creatinine | 0.83 | 1.4 | 0.9 | 3.33 | 0.9 |
| Acute stroke treatment received | No acute intervention advanced infarct progression | tPA Mechanical thrombectomy TICI 3 | Mechanical thrombectomy TICI 2C | No acute intervention patient too unstable | tPA Mechanical thrombectomy TICI 2b |
| Days from viral onset to ischemic event | 6 days | First day presentation | 14-21 days | First day presentation | 8 days |
| Pulmonary disease Severity | Severe | Severe | Mild | Severe | Severe |
| Follow-up |  |  |  |  |  |
| CT Head/MRI brain | Extensive right hemispheric infarction | No repeat imaging | Left temporal gyri, Left Basal Ganglia | Right pontine Left cerebellar | Left Parietal Post temporal Basal Ganglia |
| NIHSS | N/A | 10 | 18 | N/A | N/A |
| mRS | 6 | 6 | 4 | 6 | 5 |

HTN indicates Hypertension; DM, Diabetes Mellitus; OSA, Obstructive Sleep Apnea; MCA, Middle Cerebral Artery;
NIHSS, National Institutes of Health Stroke Scale; CXR, Chest X-ray; CT, Computer Tomography; CTA, Computer Tomography Angiography; ICA, Internal Carotid Artery; NLR, Neutrophil to Lymphocyte Ratio; TICI, Thrombolysis in Cerebral Infarction; MRI, Magnetic Resonance Imaging; mRS, modified Rankin Scale.

**Table 2:** Characteristics of Patients with Non-LVO syndromes

| Characteristic | Patient 6 | Patient 7 | Patient 8 | Patient 9 | Patient 10 |
| --- | --- | --- | --- | --- | --- |
| Demographics |  |  |  |  |  |
| Age (years) | 70-80 | 50-60 | 70-80 | 50-60 | 60-70 |
| Race | African American | Caucasian | African American | African American | African American |
| Sex | Female | Female | Female | Female | Female |
| Initial Findings |  |  |  |  |  |
| Medical history | DM, HTN | DM, HTN, ITP<br>s/p splenectomy,<br>asthma | DM, HTN | DM, HTN | DM, Asthma |
| Symptoms at disease onset | Neurological, Cough, Fever | Fever, Dyspnea | Neurological, Cough, chills | Neurological, Fever, Cough, Chills | Fever, cough, Dyspnea |
| Stroke syndrome | Multifocal emboli | Right MCA branch emboli | Lacunar | Lacunar | Left Clumsy hand/TIA |
| Cardiac Workup | NSR, No Echo | NSR, No Echo | NSR | NSR, Normal LVEF | NSR, No echo |
| Imaging |  |  |  |  |  |
| CXR | Bibasilar airspace opacities | Bilateral opacities lower fields | Right lung early infiltrate | Clear | Interstitial opacities bilateral lungs |
| CTH | No changes | Right Posterior MCA territory infarct | No changes | No changes | No changes |
| CTA Head/Neck | No LVO | No LVO | No LVO | No LVO | No LVO |
| Lab Findings |  |  |  |  |  |
| D-Dimer | 3620 | N/A | N/A | N/A | 782 |
| NLR | 10.6 | 6 | 5.1 | 5.6 | 5.1 |
| Fibrinogen | N/A | N/A | N/A | N/A | N/A |
| Ferritin | 650 | N/A | N/A | 165 | 692 |
| Creatinine | 6.08 | 0.7 | 0.63 | 1.78 | 0.67 |
| Acute Stroke treatment received | No tPA<br>No thrombectomy | No tPA<br>No thrombectomy | No tPA<br>No thrombectomy | No tPA<br>No thrombectomy | No tPA<br>No thrombectomy |
| Days since viral disease onset | 5 days | 9 days | First presentation | 5 days | 10 days |
| Pulmonary disease Severity | Severe | Mild to moderate | Moderate | Mild | Mild to Moderate |
| Post treatment/discharge |  |  |  |  |  |
| CT Head/MRI brain | Bilateral frontal/parietal multifocal infarcts | Right posterior MCA territory | Thalamocapsular infarcts | Right Internal Capsular infarct | Negative |
| NIHSS | N/A | 1 | 9 | 3 | 0 |
| mRS | 6 | 2 | 3 | 2 | 1 |

HTN indicates Hypertension; DM, Diabetes Mellitus; ITP, Idiopathic Thrombocytopenic Purpura; NIHSS, National Institutes of Health Stroke Scale; CXR, Chest X-ray; CT, Computer Tomography; CTA, Computer Tomography Angiography; NLR, Neutrophil to Lymphocyte Ratio; MRI, Magnetic Resonance Imaging; mRS, modified Rankin Scale.

The median age of large vessel stroke patients was 46 years, the youngest being in the twenties. Median NIHSS was 24. While 80% of the patients had pre-existing conditions, the most common being diabetes (70%) and hypertension (60%), no patient had known coronary disease, cerebrovascular disease, or atrial fibrillation. One of the patients developed atrial fibrillation during their hospital course. Three patients reported no prior viral symptoms at the time of presentation. The most common viral illness complaints were cough and fever. Time interval from onset of viral illness to neurological complications was 1-21 days. Two patients who presented with LVO had no prior complaints of viral illness but went on to develop a severe course of the disease.

Laboratory findings included elevated D-dimer, Neutrophil to Lymphocyte Ratio (NLR), ferritin and lactate dehydrogenase (LDH) and they correlated with severity of viral illness. Four patients also had renal involvement. One patient who presented with emergent LVO, 21 days after viral illness, was suspected to have cardiac involvement due to COVID-19. The latter patient’s transthoracic echocardiogram (TTE) showed left ventricular ejection fraction (LVEF) of 26% and global LV hypokinesis. No deep venous thromboses or pulmonary embolisms were detected in our case series.

Three of five patients who presented with large vessel occlusion (LVO) underwent thrombectomy. Two of them received tPA and had good post-thrombectomy reperfusion. Nevertheless, all but one demonstrated infarct on follow-up imaging. Most of the patients did poorly with mortality seen in three out of the five and one still critically ill. None were independent at the time of discharge. The mortality in two cases was secondary to pulmonary complications while one was due to infarct progression. The median age was 46 and NIHSS was 24. They also had a more severe form of viral illness compared with non-LVO patients. (Table 1)

Five patients presented with non-LVO syndromes. All had vascular risk factors of either diabetes or hypertension, but neither had atrial fibrillation, coronary or cerebrovascular disease. Their median age was higher (62) and NIHSS (6) lower than the LVO group. None received IV thrombolysis due to late presentation. Two had lacunar infarcts while two had infarcts related to distal embolization. One was deemed to be a transient ischemic attack (TIA). Their neurological and overall outcomes were better with only one mortality which was related to pulmonary progression. The other three were independent at the time of discharge. (Table 2)

One patient, in mid-twenties, who had no past medical history presented with NIHSS 18 and a left MCA syndrome. This patient was involved in care of COVID-19 patients in the early days of the NYC outbreak. The presentation to the ER was 8 days after known onset of viral illness while being azithromycin therapy. The patient tested positive on day 3 of the illness. Non-contrast CT Head showed early changes with ASPECTS of 7. CT angiogram confirmed a left M1 thrombus (Figure 1). At presentation, the patient was hypoxic, tachypneic and the chest x-ray (CXR) demonstrated bilateral extensive infiltrates eventually requiring intubation. IV tPA was administered within 2 hours and 15 minutes from symptom onset and patient was taken for mechanical thrombectomy. Initial cerebral angiography showed partial recanalization post-TPA and migration of the clot to distal MCA branches (Figure 1). Intra-arterial tPA was administered in those branches. There was no renal, cardiac or hepatic involvement. Initial post-operative stroke prophylaxis included ASA, clopidogrel and atorvastatin for stroke prophylaxis but switched to full dose anticoagulation with enoxaparin on day 13 of the hospitalization. The patient continues to require ventilator support due to acute respiratory distress syndrome.

**Figure 1.**
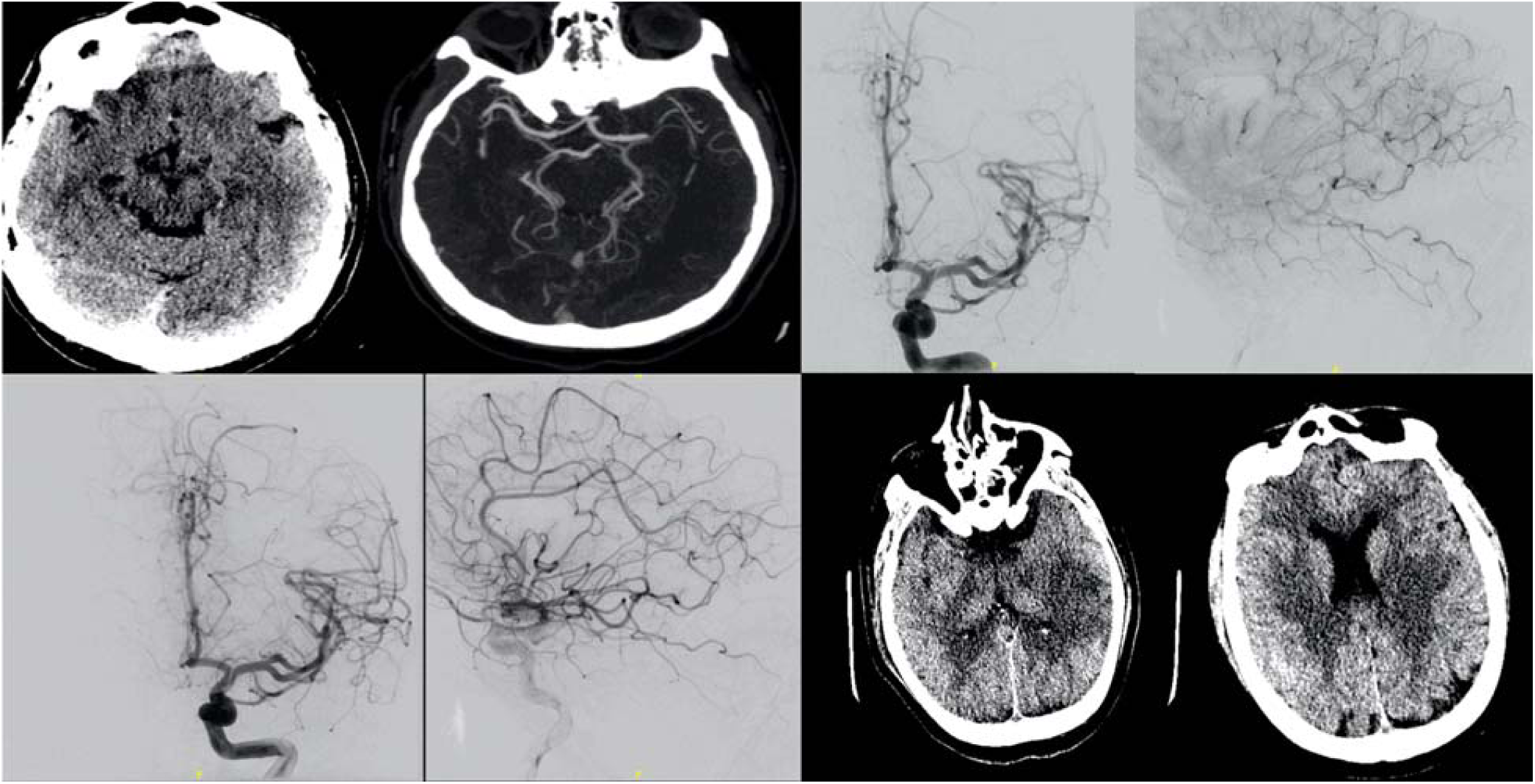
Legend: A patient with no past medical history presented to the ER after 8 days of viral illness and was on Azithromycin therapy after testing positive on day 3. NIHSS was 18 on presentation secondary to a left MCA syndrome. Non contrast CT Head is shown in the top left. Since presentation was within 4.5 hours of symptoms onset, IV tPA was administered. CTA (top left-Figure 1a) confirmed a left M1 thrombus. The patient was taken for mechanical thrombectomy and on initial cerebral angiography showed partial recanalization post-TPA and migration of the clot to distal MCA branches (top right-Figure 1b). Patient was thus given intra-arterial tPA in these branches (bottom left-Figure 1c). Post operatively infarcts in the left putamen as well as temporal and parietal regions were seen (bottom right -Figure 1d)

## Discussion

COVID-19 patients may have an asymptomatic, moderate or a severe course of illness.(10) The more severe form of the disease which requires intensive care seems to affect patients who are older, have underlying conditions and those who develop acute respiratory distress syndrome.(17) The more severe form can also affect the heart, liver, kidneys and be associated with sepsis or septic shock.(7,10)

Several case reports have also reported the possibility of a hypercoagulable condition caused by the virus.(9,11,18,19) This has specifically been seen in cases with higher incidence of VTE or PE. Although initially suspected to be due to poor prophylaxis, recent reports suggest a systemic hypercoagulable condition at play.(18,19) This is further supported by reports of autopsies which found clots in the lung, liver as well as kidneys.(10,20) This hypercoagulability is often associated with deranged lab values seen in both inflammatory conditions and consumptive coagulopathies.(9,12) These include elevation of D-dimer, Fibrin/Fibrinogen degradation products (FDP) and fibrinogen as well as shorter Thrombin Time.(9,11) Threshold of d-dimers are in fact being used as a guide by several groups to guide prophylactic systemic anticoagulation. (21)

Most neurological manifestations of COVID-19 have been non-focal presentations like headaches, encephalopathy, long tract cortical signs or seizures.(8,13,14) Li et al described the possibility of the neuro-invasiveness of SARS-COV2 being similar to other coronaviridae. They distinctly point out the possibility of a synaptic, but non-vascular, transmission of the virus to the brain.(14,22) However more severe forms of COVID-19 cases have been shown to present with strokes.(13) As per Mao et al the rate of neurovascular events in their series was about 5.7% of which about 4.9% had ischemic strokes.(8) However, direct endothelial damage mediated by the emerging role of the ACE2 receptor has neither precedent in previous coronavirus epidemics nor in sepsis-induced stroke. In addition to vascular endothelial damage, acute cardiac injury and development of antiphospholipid antibodies are important contributing factors (9).

Most ischemic stroke events in COVID-19 have been reported to be either subcortical or distal cortical distributions.(8,13) Several single case reports or small series have reported this to be concomitant in presence of other risk factors like atrial fibrillation, diabetes or hypertension.(13,16) Others have reported the possibility of several mechanisms for the stroke that maybe directly related to the infection or its complications.(8,23) These include either the causation of acute cardiac injury, creation of antiphospholipid antibodies or even the result of a severe hypercoagulable condition caused due to D-dimer or fibrinogen abnormalities.

In our series, we found that the demographic and disease features of COVID-19-related indices had a variable distribution. Our youngest patient was 27 while our oldest was 75. While the non-LVO group was older, the median age of both groups is lower than would be expected for the general population. In the case of LVO, a median age of 48 is expressly unusual. Only 40% of our cases were male which is different from the typical male preponderance in other studies. D-dimer was available in 70% of the cases and levels varied from 391 to 32000. Almost 80% had pre-existing conditions like diabetes and hypertension though none of them had history of atrial fibrillation or were chronic smokers. We observed that only six patients (60%) sought emergent medical care due to stroke symptoms, which result in a delay in care. Only three patients had a severe course of the pulmonary disease prior to the neurological event requiring them to be hospitalized. One was asymptomatic for three weeks. Five patients presented with emergent large vessel occlusion of which three underwent thrombectomy. One post-thrombectomy patient died from pulmonary progression. Most LVO cases did poorly with three ending in mortality while another one in a critical condition due to their lungs. All of these had high neutrophil/lymphocyte ratio except the one who had some neurological recovery. The LVO cases were typically younger, had a worse neurological presentation, more severe form of viral disease and higher levels of hypercoagulability markers than the non-LVO patients.

In terms of our stroke etiological distributions, we encountered the gamut of small vessel strokes, branch or multifocal emboli, TIAs and large vessel occlusions. Most of these patients had vascular risk factors but nearly a fifth did not, and none had known coronary or cerebrovascular disease. Thus, it is possible that COVID-19 related thromboembolism may follow multiple separate pathogenetic pathways. In patients with pre-existing vascular risk factors, it may predispose to the ischemic event earlier than if purely driven by those underlying conditions. This could explain why some of our younger patients in the 40-50 age range presented with stroke. In these situations, it may follow the paradigm seen in other hypercoagulable conditions where two or more factors act synergistically to increase risk of stroke.(12) It can also be likened to the double-hit theory of many cancers.(24) It is also possible that in some cases this hypercoagulable condition by itself is enough to cause an embolic stroke. This may be correlated with elevated lab markers like D-Dimer or NLR ratio or presence of antiphospholipid antibodies.(8,23) A third pathway maybe related to its effect on the heart when it causes cardiomyopathy or myocarditis which in turn may predispose to cardio-embolism.(16,25) This mechanism may be mediated by ACE2 targeting which could affect the vascular endothelium or the heart directly. Finally, since the incidence of VTE is higher in these cases, the possibility of paradoxical emboli cannot be ignored.(12) To our knowledge, this is the first study that attempts to define the ischemic stroke etiologic subtype as well as pathophysiology, direct or indirect, in patients with SARS-COV-2 Virus.

### Limitations

Our study has several limitations. This is a short analysis of neurovascular cases presenting to the hospital. As the number of cases in New York City and our hospitals continue to accumulate, we cannot determine the true rate of neurovascular events in COVID patients. Due to the stress on the resources created by COVID-19 some of the patients did not receive advanced imaging such as MRI to confirm whether a silent infarct may have occurred. Additionally, many cases had other risk factors and the embolic events may have been related to the underlying risk factor rather than the viral infection, or simply exacerbated by it. Finally, the type of strokes caused by the disease fit into several etiologic pathway rendering it difficult to delineate an appropriate treatment or prevention strategy.

## Conclusion

Stroke in the setting of COVID-19 has an unusual presentation including atypical demographics and delayed time windows. Ischemic events can be small vessel, branch emboli or large vessel occlusions. The latter is often associated with either a hypercoagulable state or cardio-embolism. Ischemic stroke can be a presenting symptom of COVID-19 and may not always be associated with severe disease markers including in the young, minorities and healthcare workers.

## Data Availability

all data owned by authors and can be shared upon appropriate request via correspondent author

## Notes

### Competing Interest Statement

The authors have declared no competing interest.

### Funding Statement

no funding

